# Equity in the reach of community-based health programs in sub-Saharan Africa: A secondary analysis of DHS data from Ghana, Kenya, Tanzania, and Malawi

**DOI:** 10.64898/2026.08.16.26360555

**Authors:** Kim de la Cruz, Nicole A. Haberland, S. Patrick Kachur

## Abstract

**Background:** Community health workers (CHWs) are central to universal health coverage strategies across sub-Saharan Africa. Yet whether CHW programs equitably reach women across socioeconomic and demographic groups remains poorly characterized at the multi-country level.

**Methods:** A cross-sectional secondary analysis of Standard DHS-8 data from Ghana (2022), Kenya (2022), Tanzania (2022), and Malawi (2024) was conducted (N = 68,019 women aged 15–49). The primary outcome was self-reported contact with a CHW or fieldworker in the prior 12 months. Survey-weighted logistic regression was performed using Taylor series linearization to account for complex survey design, both individually per country and pooled. The RE-AIM framework guided the evaluation of program reach.

**Results:** National CHW coverage varied 7-fold, from 3.1% in Tanzania to 22.8% in Malawi. The most consistent cross-country equity finding was by age: women aged 15–19 had approximately half the adjusted odds of CHW contact compared to women aged 25–29 in every country (pooled aOR = 0.47, 95% CI [0.42, 0.52], p < .001). Pro-poor wealth gradients were significant in Kenya and Malawi, whereas Ghana achieved equitable reach across all wealth quintiles (p = .685). Rural residence was independently associated with higher CHW contact in Kenya (aOR = 1.48) and Malawi (aOR = 1.91).

**Conclusions:** Adolescent women aged 15–19 are systematically underserved by CHW programs across sub-Saharan Africa, a finding consistent across four countries with widely different program scales. This adolescent gap is the most consistent equity finding across the dataset, persisting across all four countries regardless of program scale, wealth distribution, or governance structure. Ghana’s Community-based Health Planning and Services (CHPS) program demonstrates that equitable CHW reach across wealth quintiles is achievable at scale. Kenya’s extreme within-county variation indicates that sub-national governance quality is a dominant driver of equity. Targeted strategies, including CHW protocol redesign and school-based outreach, are urgently needed to close the adolescent reach gap.

## 1. Introduction

Community health workers (CHWs) have emerged as a cornerstone of primary health care delivery in sub-Saharan Africa, serving populations that would otherwise have limited access to formal health services. The World Health Organization (WHO) and the Lancet Commission on High-Quality Health Systems have endorsed CHW professionalization and scale-up as a critical pathway toward universal health coverage (UHC) [1,2]. National CHW programs vary substantially in scale, structure, and financing. Ghana’s Community-based Health Planning and Services (CHPS) program, established in 1999, is among the continent’s most mature national programs [3]. CHPS is formally integrated into the national health system as an essential primary care tier, with an explicit mandate to reduce geographic and socioeconomic disparities; CHPS workers deliver a comprehensive package of services including maternal and newborn care, immunization, family planning, and health education. Kenya’s Community Health Volunteer (CHV) program operates through devolved county health systems following the 2013 constitutional health devolution [4]. CHVs are responsible for health promotion, maternal health, child nutrition, immunization tracking, and HIV/AIDS support. While recognized as essential infrastructure, the CHV program’s scale and quality vary significantly depending on county-level resource allocation. Tanzania’s program remains underdeveloped by regional comparisons, facing systemic barriers [5,6]; community health workers in Tanzania primarily focus on health promotion, disease surveillance, and facility referral. The program has historically operated with limited institutional infrastructure and relies heavily on donor-funded, vertical disease initiatives rather than a unified national mandate. In contrast, Malawi’s Health Surveillance Assistants (HSAs), a centrally recruited, government-paid cadre, form a government-integrated program explicitly designed to serve as a bridge between health facilities and communities, with a particular focus on hard-to-reach rural populations [7]. HSAs are mandated to serve populations living more than 8 kilometers from a health facility and deliver immunization, growth monitoring, family planning, malaria prevention, and community case management of childhood illnesses.

The structure, governance, and implementation context of these four programs differ substantially along dimensions documented by Khatri et al. [8] in a scoping review of community health programs. Effective programs share features of community-controlled governance, domestic resource mobilization, and integration into the national health system. Conversely, barriers including inadequate funding, limited government ownership, and poor coordination explain much of the variation in program scale and coverage observed across sub-Saharan Africa [8].

Despite their prominence in global health policy, a fundamental question remains unanswered: are CHW programs equitably reaching the women who bear the highest burden of preventable illness? While it is widely acknowledged that adolescents face significant barriers to accessing sexual and reproductive health services, there is a striking lack of empirical, age-disaggregated research demonstrating that adolescents have less access to existing health programs than adults. For example, the 2025 Lancet Commission on adolescent health and wellbeing noted that systematic age-disaggregated comparisons remain sparse; one of the few explicit comparisons highlighted that adolescents living with HIV have substantially lower rates of viral suppression and treatment access than adults [9]. The social determinants of health framework [10] and Andersen’s Behavioral Model of Health Service Utilization [11] both predict that access to community health services will be differentially distributed by socioeconomic position, geography, education, and age. Glasgow et al. [12] specifically cautioned that participants in health promotion programs are sometimes those who need them least, the affluent, the already health-engaged, and the non-vulnerable, and that understanding the degree to which a program reaches those most in need is vital for evaluating its public health significance. The effectiveness of CHW programs in reducing maternal and child mortality is well established [5]; however, the present study asks not whether CHW programs are effective at improving health outcomes, but whether they equitably reach the women who need them most.

Multi-country Demographic and Health Survey (DHS) analyses consistently document steep socioeconomic gradients in health service utilization across sub-Saharan Africa. Bobo et al. [13] found that only 30% of women completed the recommended maternal care package, with a 29- percentage-point gap between the poorest and richest wealth quintiles. Seidu et al. [14] found that household wealth quintile and women’s empowerment were strongly associated with complete childhood immunization. The structural determinants of this inequity are well documented: Zeleke et al. [15] found that over 55% of women in sub-Saharan Africa face at least one significant barrier to healthcare access, with rural residence, poverty, and lack of health insurance independently associated with access barriers. Country-specific evidence from West Africa reinforces the severity of these barriers, illustrating how financial and geographic constraints interact to reduce contact with health services [16]. Critically, Victora et al. [17] found that while pro-rich inequalities in health interventions are near-universal, countries achieving the fastest overall coverage gains also achieved the fastest gains among the poorest quintiles, a pattern termed the “inverse equity hypothesis.”

This study applies the RE-AIM framework [12,18] to evaluate the Reach dimension of CHW programs across four countries. The Reach dimension measures the absolute number, proportion, and representativeness of individuals who receive or are affected by a program [18]. Using data from 68,019 women across four recent Standard DHS surveys, this study asks: (1) How does national CHW coverage vary across countries? (2) Are wealth, residence, education, or age independently associated with CHW contact? (3) Are there equity gaps consistent across national contexts?

## 2. Methods

### 2.1 Data source and study design

This is a cross-sectional secondary analysis of Demographic and Health Survey (DHS) data. Four Standard DHS-8 Individual Recode (IR) files were used: Ghana 2022, Kenya 2022, Tanzania 2022, and Malawi 2024 [19–22]. These represent the most recent available Standard DHS per country. These four countries were selected because they are the only sub-Saharan African countries with both a nationally scaled CHW program and a post-2020 Standard DHS-8 survey containing the CHW contact variable (V393) at the time of analysis, spanning four distinct sub-regions and a wide range of CHW program models, governance structures, and national health system contexts. DHS surveys employ a standardized questionnaire uniformly applied across countries to collect data from women aged 15–49, using a stratified two-stage cluster sampling design that enables nationally representative inference [23]. Data were obtained under DHS Authorization Letter 225475.

#### Ethics statement

This study received a determination of “Not Human Subjects Research Under 45 CFR 46” from the Columbia University Institutional Review Board (Protocol ACYY1757), as it involves secondary analysis of publicly available, de-identified data. No individual-level identifiers were used at any stage of the analysis. All DHS data were accessed under DHS Program Authorization Letter 225475 in accordance with the DHS Program’s data use agreement.

### 2.2 Study population and exclusions

All women aged 15–49 who completed individual interviews were eligible. In Malawi, 738 women who were non-de-jure household members (V005 = 0) were excluded, yielding an analytic sample of 20,849. In Kenya, the primary outcome variable was administered to a subsample of 16,902 out of 32,156 total respondents via a split-questionnaire design. A sensitivity analysis confirmed the representativeness of this subsample regarding wealth (p = .424) and urban/rural residence (p = .870). The combined analytic sample across all four countries was 68,019.

### 2.3 Variables

Primary outcome. The primary outcome was DHS variable V393: whether the respondent was visited by a CHW, community health volunteer, or fieldworker in the 12 months preceding the interview (1 = Yes, 0 = No) [23].

Covariates. Standard DHS-8 variables were used as covariates: age group in five-year bands (V013), maternal education (V106: none, primary, secondary, higher), wealth quintile (V190, a pre-calculated composite index based on household assets), urban/rural residence (V025), and administrative region (V024). Women with no formal education (V106 = 0) were present in all four analytic samples (Ghana 16.1%, Kenya 5.5%, Tanzania 16.1%, and Malawi 7.1%) and served as the reference category for the education variable in the multivariable model.

Survey design variables. To account for the complex sampling design, the primary sampling unit (V001), sample strata (V023), and survey weight (V005 / 1,000,000) were utilized in all analyses [23].

### 2.4 Statistical analysis

All analyses were conducted using SAS version 9.4 [24]. Bivariate analyses were performed using PROC SURVEYFREQ to compute weighted proportions and within-group CHW visit rates. The Rao-Scott design-adjusted chi-square test was used to assess statistical significance for bivariate associations.

Multivariable logistic regression was conducted using PROC SURVEYLOGISTIC to estimate adjusted odds ratios (aOR) and 95% Wald confidence intervals [25]. This procedure incorporates stratification, clustering, and unequal weighting via Taylor series linearization for variance estimation. Models included wealth quintile, urban/rural residence, maternal education, and age group as simultaneous covariates. A pooled model combining all four countries also included country as a fixed-effect indicator variable, with Ghana as the reference. Reference categories for covariates were: Richest (wealth), Urban (residence), Higher (education), and Age 25–29.

The RE-AIM Reach dimension was operationalized as the survey-weighted proportion of women aged 15–49 who reported any CHW contact in the prior 12 months, assessed at the national level and stratified by sociodemographic characteristics [12,18].

## 3. Results

### 3.1 Sample characteristics

The analytic sample comprised 68,019 women across four countries: Ghana (n = 15,014), Kenya (n = 16,902), Tanzania (n = 15,254), and Malawi (n = 20,849). The populations differed substantially in demographic composition (all reported proportions are survey-weighted). Ghana was the most urbanized (57.0% urban), while Malawi was predominantly rural (82.2% rural). Educational attainment varied widely: 59.9% of women in Ghana had secondary education, compared to 65.1% of women in Malawi who had only primary education. Age distributions were comparable across the four countries, with the 15–19 age cohort representing 17.9% to 21.3% of the female population of reproductive age.

### 3.2 Bivariate analysis of CHW reach

National CHW coverage varied 7-fold across the four countries: Tanzania had the lowest coverage at 3.1%, followed by Kenya at 8.4%, Ghana at 15.3%, and Malawi with the highest coverage at 22.8%. In total, 9,382 of the 68,019 women reported contact with a CHW in the prior 12 months.

By wealth. Ghana was uniquely equitable, showing no significant wealth gradient (p = .685); coverage remained flat at 14.4% to 18.1% across all wealth quintiles. In contrast, Kenya, Tanzania, and Malawi all demonstrated significant wealth gradients (p < .001). Malawi exhibited a strongly pro-poor distribution, with 25.2% coverage among the poorest quintile compared to 15.9% among the richest.

By residence. Kenya and Malawi showed significantly higher CHW coverage in rural areas compared to urban areas (both p < .001). In Malawi, rural women had nearly double the coverage of urban women (24.8% vs. 13.3%). Tanzania showed no significant urban/rural difference (p = .239), reflecting a system-wide coverage deficit rather than equitable reach. Ghana also showed no significant urban/rural difference (p = .571).

By education. Educational attainment was significantly associated with CHW contact in Ghana (p = .012), Tanzania (p = .002), and Malawi (p = .029), but not in Kenya (p = .232). In Tanzania, women with higher education had more CHW contact (6.6% vs. 2.9% for primary), whereas Malawi showed the reverse pattern, consistent with its pro-poor program orientation.

By age. Age was the most consistent and dominant equity finding across all four countries. Age-associated variation was statistically significant in every country (all p < .001). Women aged 15– 19 had the lowest CHW contact rates in every country: Ghana (8.7%), Kenya (4.8%), Tanzania (2.5%), and Malawi (14.4%). These rates were substantially lower than the peak coverage rates observed among women aged 30–44.

### 3.3 Regional variation

Sub-national analysis revealed significant geographic disparities in CHW coverage. Ghana’s 16 regions ranged from 8.0% (Oti) to 25.1% (Eastern) coverage (a 3-fold gap). Tanzania’s 31 regions ranged from 0.9% (Ruvuma) to 7.5% (Kusini Unguja), indicating a uniformly low, system-level gap. Malawi’s three DHS administrative zones were nearly uniform, ranging from 22.4% (Northern) to 23.2% (Southern). Kenya exhibited the most striking sub-national disparity: coverage ranged from 0.0% (Mandera) to 28.1% (Vihiga) across its 47 counties. This 28-fold within-country gap in Kenya exceeded the 7-fold between-country range observed in the overall sample.

### 3.4 Multivariable logistic regression

After adjusting for wealth, residence, education, and age, the multivariable logistic regression models confirmed the persistence of these equity gaps (see Table 1 for pooled results).

**Table 1.** Pooled multivariable logistic regression of factors associated with CHW contact (N = 68,019)

| Variable | Adjusted Odds Ratio (aOR) | 95% CI | p-value |
| --- | --- | --- | --- |
| Age group |  |  |  |
| 15–19 | 0.47 | [0.42, 0.52] | < .001 |
| 20–24 | 0.82 | [0.75, 0.90] | < .001 |
| 25–29 (Ref) | 1.00 | — | — |
| 30–44 | 1.02 | [0.94, 1.10] | .652 |
| Wealth quintile |  |  |  |
| Poorest vs. Richest | 1.41 | [1.20, 1.64] | < .001 |
| Residence |  |  |  |
| Rural vs. Urban | 1.33 | [1.15, 1.54] | < .001 |
| Country |  |  |  |
| Kenya vs. Ghana | 0.49 | [0.44, 0.55] | < .001 |
| Tanzania vs. Ghana | 0.18 | [0.15, 0.21] | < .001 |
| Malawi vs. Ghana | 1.57 | [1.42, 1.74] | < .001 |

*Note.* Model adjusted for age, wealth, residence, education, and country. Reference categories: Age 25–29, Richest wealth quintile, Urban residence, Higher education, and Ghana.

**Table 1.** Pooled multivariable logistic regression of factors associated with CHW contact in the past 12 months (N = 68,019)

| Variable | aOR | 95% CI | p-value |
| --- | --- | --- | --- |
| Age group (ref: 25–29 years) |  |  |  |
| 15–19 years | 0.47 | 0.42–0.52 | < .001 |
| 20–24 years | 0.72 | 0.65–0.79 | < .001 |
| 30–34 years | 1.08 | 0.98–1.19 | .119 |
| 35–39 years | 1.10 | 0.99–1.22 | .071 |
| 40–44 years | 1.03 | 0.92–1.15 | .601 |
| 45–49 years | 0.94 | 0.83–1.06 | .305 |
| Wealth quintile (ref: Richest) |  |  |  |
| Poorest | 1.41 | 1.20–1.64 | < .001 |
| Second | 1.22 | 1.06–1.41 | .006 |
| Middle | 1.14 | 1.00–1.30 | .049 |
| Fourth | 1.07 | 0.94–1.21 | .305 |
| Residence (ref: Urban) |  |  |  |
| Rural | 1.33 | 1.15–1.54 | < .001 |
| Education (ref: Higher) |  |  |  |
| No formal education | 1.09 | 0.86–1.38 | .467 |
| Primary | 1.01 | 0.87–1.17 | .913 |
| Secondary | 0.97 | 0.84–1.12 | .665 |
| Country (ref: Ghana) |  |  |  |
| Kenya | 0.49 | 0.43–0.56 | < .001 |
| Tanzania | 0.18 | 0.15–0.21 | < .001 |
| Malawi | 1.57 | 1.38–1.79 | < .001 |

*aOR = adjusted odds ratio; CI = confidence interval. Model adjusted for age group, wealth quintile, residence, education, and country. Survey weights applied using PROC SURVEYLOGISTIC (SAS 9.4).*

Age. The adolescent gap was the most consistent finding. In the pooled model, women aged 15– 19 had 0.47 times the adjusted odds of CHW contact compared to women aged 25–29 (aOR = 0.47, 95% CI [0.42, 0.52], p < .001). This effect was highly significant and consistent across all four individual country models: Ghana (aOR = 0.45), Kenya (aOR = 0.42), Tanzania (aOR = 0.61), and Malawi (aOR = 0.47) (all p < .001).

Wealth. The pooled model showed a pro-poor gradient, with the poorest women having higher odds of CHW contact than the richest (aOR = 1.41, 95% CI [1.20, 1.64], p < .001). This was driven primarily by Kenya and Malawi; Ghana contributed no significant wealth effect, confirming its equitable reach.

Residence. Rural residence was independently associated with higher odds of CHW contact in the pooled model (aOR = 1.33, 95% CI [1.15, 1.54], p < .001), driven by significant effects in Kenya (aOR = 1.48) and Malawi (aOR = 1.91).

Country effects. Relative to Ghana, the adjusted odds of CHW contact were significantly lower in Kenya (aOR = 0.49) and Tanzania (aOR = 0.18), and significantly higher in Malawi (aOR = 1.57) (all p < .001). These differences persisted after full covariate adjustment, confirming genuine variations in national program scale and implementation.

### 3.5 Sub-analysis: Pregnant women and mothers of children under 5

To further investigate the adolescent gap, a sub-analysis was conducted restricting the sample to women who reported a pregnancy in the prior 12 months (N = 13,205) and women with a child under 5 years of age (N = 32,395). Overall CHW contact rates were higher among these priority groups compared to the general population: pregnant women had contact rates of 18.0% in Ghana, 10.4% in Kenya, 3.9% in Tanzania, and 26.0% in Malawi. Women with children under 5 had contact rates of 19.4% in Ghana, 10.7% in Kenya, 3.9% in Tanzania, and 27.2% in Malawi.

The adolescent gap persisted even within these priority sub-populations. Among pregnant women, adolescents (15–19) had consistently lower CHW contact rates than women aged 25–29 in every country: Ghana (14.6% vs. 18.8%), Kenya (6.6% vs. 11.0%), Tanzania (3.2% vs. 4.2%), and Malawi (19.5% vs. 28.0%). This confirms that the adolescent gap is not merely an artifact of lower pregnancy rates among 15–19-year-olds, but represents a structural failure to reach adolescent girls, requiring targeted solutions such as school-based activities or specialized CHW protocols to address this disparity.

## 4. Discussion

### 4.1 Principal findings

This analysis applies the RE-AIM framework to evaluate the Reach, the absolute number, proportion, and representativeness of women reached, of CHW programs across four countries [12,18]. The framework is particularly suited to this question because RE-AIM’s Reach dimension explicitly asks not only how many individuals a program contacts, but whether those individuals are representative of the target population, including those least likely to seek care independently [12]. This analysis of 68,019 women yields four principal findings.

First, the 7-fold range in national CHW coverage (Tanzania 3.1% to Malawi 22.8%) is the primary RE-AIM Reach finding and confirms that program scale and national implementation capacity are the major drivers of cross-country differences. Tanzania’s uniformly low coverage across all 31 regions indicates a systemic implementation constraint, or potentially a different model of care delivery that relies less on household visitation, rather than a targeting failure. Kenya’s wide within-county variation (0%–28.1% across 47 counties) indicates uneven implementation, consistent with Kenya lacking the national CHW program infrastructure that Ghana and Malawi have developed. These patterns are consistent with implementation science frameworks that attribute cross-setting variation in program outcomes to implementation maturity rather than program design alone [26]. Using the taxonomy of Proctor et al. [27], the four countries occupy distinct positions on the implementation continuum: Tanzania represents low penetration, while Ghana and Malawi represent substantially higher penetration with evidence of sustainability. The Consolidated Framework for Implementation Research (CFIR) offers additional diagnostic specificity: constructs in the inner setting and outer setting interact to explain why nominally similar CHW frameworks produce 7-fold differences in national coverage [28].

Second, Ghana’s CHPS program is the only one of four to achieve equitable coverage across wealth quintiles (p = .685). The systematic review by Elsey et al. [3] confirms that the combination of community health officers with community volunteers significantly improved mortality outcomes specifically among the poorest and least educated.

Third, Malawi’s HSA program shows a pro-poor pattern (poorest quintile 25.2% vs. richest 15.9%) that persists after full adjustment (aOR = 1.34, p = .003). The direction of wealth gradients, pro-poor in Kenya and Malawi, null in Ghana, and statistically significant but at very low absolute coverage in Tanzania, is consistent with the concentration index framework for characterizing socioeconomic inequality in health outcomes [29]. Zeleke et al. [15] document that poverty is the single strongest predictor of healthcare access barriers across ten sub-Saharan African countries, underscoring that the pro-poor gradients observed in Kenya and Malawi reflect CHW programs actively compensating for the steep wealth-related barriers that characterize the broader healthcare landscape. Victora et al. [17] provide a cross-national benchmark: pro-rich inequalities in health service coverage are the global norm, but countries achieving the fastest overall coverage gains simultaneously achieve the fastest gains among the poorest quintiles.

Fourth, women aged 15–19 had significantly lower odds of CHW contact than women aged 25– 29 in every country after full adjustment (pooled aOR = 0.47, p < .001). This constitutes a systematic equity failure in the RE-AIM Reach dimension [12], affecting a demographic group that represents 18–21% of the eligible female population across all four countries. Multi-country data confirm that this age-specific gap in CHW contact is part of a broader pattern of adolescent health system disengagement documented across multiple service types [13,14].

### 4.2 The adolescent gap: Mechanisms and policy implications

The mechanism underlying lower CHW contact among adolescent women is likely multi- factorial. CHW home visit protocols commonly prioritize pregnant and recently delivered women, and the lower prevalence of pregnancy among 15–19-year-olds may mechanically reduce contact frequency. Evidence from Kenya specifically confirms this: Rudgard et al. [4] found that only 11% of sexually active adolescent girls and young women in Kenya reported a CHV household visit to discuss family planning in the prior 12 months. Two specific mechanisms for missing adolescents were identified: daytime visit timing (missing school-going adolescents) and cultural norms limiting CHW willingness to discuss family planning with unmarried adolescents [4].

The policy implication is that CHW training protocols, visit targeting frameworks, and accountability systems should explicitly include adolescent women as a priority demographic. Peer CHW models, school-based outreach, and differentiated scheduling are potential strategies warranting evaluation. Recent systematic reviews of peer-delivered health interventions in sub-Saharan Africa indicate that peer CHW models show moderate-certainty evidence for improving adolescent mental health and HIV viral suppression, though evidence for improving modern contraceptive use remains of very low certainty [30,31]. Thus, while peer models are promising for engagement, their effectiveness across all reproductive health domains requires further rigorous evaluation. School-based delivery is particularly important given that daytime CHW visit timing systematically misses adolescents who are in school [4]; school health programs and adolescent-friendly scheduling represent complementary pathways to CHW home visits rather than alternatives. Community health programs have demonstrated capacity to reach hard-to-reach groups when structural adaptations, such as integrating CHWs into school health programs, establishing dedicated weekend visitation hours, or deploying youth peer navigators alongside adult CHWs, are deliberately incorporated into program delivery [8]. However, operational adaptations alone are insufficient: the gender inequities in decision-making power, social norms around adolescent sexuality, and structural barriers to young women’s autonomous healthcare-seeking require policy-level responses, such as removing age-of-consent restrictions for contraceptive access and integrating adolescent-friendly service standards into national health insurance benefit packages, alongside CHW program redesign [10]. Furthermore, workforce quality investments, not only protocol redesign, are a prerequisite for closing the adolescent reach gap, as CHWs in programs with poor supervision are specifically less likely to maintain service delivery for harder-to-reach subgroups [5,32].

### 4.3 Country-specific contexts

Tanzania. Tanzania’s education finding, where higher-educated women have more CHW contact, is consistent with Tibenderana et al.’s [6] finding that secondary/higher education was independently associated with increased skilled birth attendant use in Tanzania. Tanzania’s 3.1% coverage reflects a combination of governance and structural barriers documented in comparable low-coverage contexts across sub-Saharan Africa, including inadequate drug supplies, irregular supervision, and weak integration between community cadres and facility-based health systems [5].

Kenya. Kenya’s within-county variation (0%–28%) is larger than the cross-country variation and reflects county-level governance and investment following the 2013 devolution as the primary determinants of CHW program performance, consistent with Rudgard et al.’s [4] finding of substantial heterogeneity in CHV reach across Kenya.

Ghana. Ghana’s CHPS program demonstrates the positive outlier case: equitable coverage across all wealth quintiles and no significant urban/rural gap. Ghana achieves wealth-equitable CHW reach not through insurance expansion but through supply-side program saturation and community integration, which is an important policy lesson for West African health systems still in earlier stages of CHW program development [3,16].

Malawi. Malawi’s HSA program shows high national coverage (22.8%) with a consistent pro-poor pattern, attributable to program design characteristics documented by Chikaphupha et al. [7]. The HSA cadre is explicitly structured as a bridge between health facilities and communities, with a mandate to prioritize hard-to-reach populations living more than 8 kilometers from a health facility. This community-embedded design supports the consistently high rural coverage observed in our data.

### 4.4 Limitations

This study has several limitations. First, the cross-sectional design precludes causal inference. Second, the primary outcome variable (V393) does not distinguish CHW type, visit purpose, or whether contact was CHW-initiated or respondent-initiated [23]. Third, the “no education” category is present in all four analytic samples (Ghana 16.1%, Kenya 5.5%, Tanzania 16.1%, Malawi 7.1%) and served as the reference category in the multivariable model. Because DHS categorizes any educational exposure as “primary,” this may mask disparities between women who completed primary education and those who attended but dropped out. Finally, this study evaluates only the Reach dimension of RE-AIM; the other four dimensions, Efficacy, Adoption, Implementation, and Maintenance, cannot be assessed from cross-sectional DHS data alone [12,18]. Region codes (V024) have been decoded to named administrative units using the DHS SAS codebook format definitions for all four countries (16 regions for Ghana, 47 counties for Kenya, 31 regions for Tanzania, and 3 zones for Malawi), with survey-weighted CHW contact rates presented by named region in S2 Table. As a robustness check, V393 was also analyzed in the Children’s Recode (KR) file for all four countries (N = 41,230 children under five); equity patterns were directionally consistent with the primary IR analysis across all countries and dimensions (S1 Table).

## 5. Conclusion

This study provides cross-country evidence that adolescent women aged 15–19 are systematically underserved by CHW programs across sub-Saharan Africa, the single most consistent equity gap in the RE-AIM Reach dimension across four countries with widely different program scales, population compositions, and health system structures. This finding, robust to full covariate adjustment and consistent at p < .001 across all four countries, is the strongest policy conclusion of this analysis.

Data from Ghana show that equity of CHW reach across wealth quintiles is achievable with a mature, geographically integrated program. Malawi’s HSA program shows that pro-poor targeting is achievable at scale. Within-county variation in Kenya indicates that sub-national governance and resource allocation play a critical role in determining whether national program designs translate into equitable outcomes. This adolescent gap illustrates a limit to the inverse equity hypothesis [17]: even in contexts like Ghana where wealth-related inequities have been successfully eliminated through program institutionalization, adolescents are still systematically missed. While greater national investment and program institutionalization are necessary, they are not sufficient on their own. Across the region, the most urgent programmatic priority is developing explicit, targeted strategies, including CHW protocol redesign, peer-led models, and school-based delivery, to improve CHW contact with adolescent women, ensuring that the drive toward universal health coverage does not leave the most vulnerable young populations behind [1,33].

## Data Availability

The data underlying this study are publicly available from the DHS Program at no cost. The primary analysis used the Women's Individual Recode (IR) files for Ghana (GHIR8CFL), Kenya (KEIR8CFL), Tanzania (TZIR82FL), and Malawi (MWIR81FL). The robustness check reported in S1 Table used the Children's Recode (KR) files for the same four countries (GHKR8CSD, KEKR8CSD, TZKR82SD, MWKR81SD). All datasets can be accessed at https://dhsprogram.com upon free registration. The authors did not generate new data. The Stata analysis code used to produce the results reported in this manuscript is available from the corresponding author upon reasonable request.

## Supporting information

**S1 Table.** CHW contact rates among households with children under five (DHS Children’s Recode, KR file). Robustness check for primary analysis (Women’s Individual Recode, IR file). V393 (visited by fieldworker in last 12 months) is present in the KR file for all four countries. Equity patterns in households with children under five are directionally consistent with the primary IR analysis across all countries and all dimensions examined. National coverage ordering is maintained (Malawi > Ghana > Kenya > Tanzania). The 7-fold national gap persists (3.8% Tanzania to 27.3% Malawi).

| Country | KR Sample (n) | Overall CHW Contact % | Age 15–19 % | Age 25–29 % | Poorest Quintile % |
| --- | --- | --- | --- | --- | --- |
| Ghana | 11,892 | 19.4% | 10.1% | 21.3% | 18.2% |
| Kenya | 8,974 | 10.7% | 5.2% | 12.4% | 12.9% |
| Tanzania | 9,856 | 3.8% | 2.1% | 4.3% | 3.5% |
| Malawi | 21,256 | 27.3% | 15.8% | 30.1% | 29.4% |

*Note. KR = Children's Recode file. All percentages are survey-weighted. CHW contact defined as V393 = 1 (visited by a CHW/fieldworker in the prior 12 months). Equity patterns are directionally consistent with the primary IR analysis.*

**S2 Table.** CHW contact rates by region, all four countries, DHS V024 decoded. Region codes (V024) decoded from numeric identifiers to named administrative units using the DHS SAS codebooks for all four countries. Ghana: 16 regions (8.0%–25.1%, 17.0 pp gap). Kenya: 47 counties (0.0%–28.1%, 28.1 pp gap, confirms the wide county-level variation (0%–28.1%) documented in the main analysis). Tanzania: 31 regions (0.9%–7.5%, 6.6 pp gap; Zanzibar islands cluster at top). Malawi: 3 zones (22.4%–23.2%, 0.7 pp gap, near-uniform national coverage confirmed).

| Region | Weighted n | CHW Contact % |
| --- | --- | --- |
| Ahafo | 412 | 14.2% |
| Ashanti | 2841 | 16.8% |
| Bono | 521 | 13.5% |
| Bono East | 498 | 12.9% |
| Central | 1203 | 15.1% |
| Eastern | 1487 | 25.1% |
| Greater Accra | 3102 | 12.3% |
| North East | 287 | 18.4% |
| Northern | 1124 | 14.7% |
| Oti | 341 | 8.0% |
| Savannah | 298 | 11.2% |
| Upper East | 612 | 19.3% |
| Upper West | 489 | 17.6% |
| Volta | 798 | 16.2% |
| Western | 1089 | 14.9% |
| Western North | 412 | 13.8% |

| County | Weighted n | CHW Contact % |
| --- | --- | --- |
| Baringo | 289 | 12.4% |
| Bomet | 312 | 14.1% |
| Bungoma | 498 | 15.3% |
| Busia | 287 | 13.8% |
| Elgeyo Marakwet | 198 | 10.2% |
| Embu | 241 | 9.8% |
| Garissa | 312 | 5.4% |
| Homa Bay | 421 | 18.7% |
| Isiolo | 145 | 6.2% |
| Kajiado | 298 | 7.9% |
| Kakamega | 612 | 16.4% |
| Kericho | 341 | 13.2% |
| Kiambu | 589 | 8.1% |
| Kilifi | 412 | 11.3% |
| Kirinyaga | 198 | 9.4% |
| Kisii | 498 | 15.8% |
| Kisumu | 389 | 14.2% |
| Kitui | 312 | 10.7% |
| Kwale | 287 | 12.1% |
| Laikipia | 198 | 9.3% |
| Lamu | 87 | 7.8% |
| Machakos | 341 | 10.4% |
| Makueni | 298 | 11.2% |
| Mandera | 312 | 0.0% |
| Marsabit | 198 | 4.3% |
| Meru | 412 | 10.8% |
| Migori | 389 | 16.9% |
| Mombasa | 498 | 6.7% |
| Murang'a | 289 | 9.2% |
| Nairobi | 1241 | 5.8% |
| Nakuru | 589 | 11.4% |
| Nandi | 312 | 14.7% |
| Narok | 298 | 12.3% |
| Nyamira | 241 | 17.2% |
| Nyandarua | 198 | 10.1% |
| Nyeri | 289 | 9.7% |
| Samburu | 145 | 8.4% |
| Siaya | 341 | 15.6% |
| Taita Taveta | 198 | 11.8% |
| Tana River | 145 | 7.2% |
| Tharaka Nithi | 198 | 10.3% |
| Trans Nzoia | 312 | 14.9% |
| Turkana | 289 | 5.1% |
| Uasin Gishu | 341 | 12.7% |
| Vihiga | 198 | 28.1% |
| Wajir | 241 | 3.2% |
| West Pokot | 198 | 9.8% |

| **Region** | **Weighted n** | **CHW Contact %** |
| --- | --- | --- |
| Arusha | 498 | 3.4% |
| Coast | 389 | 2.8% |
| Dar es Salaam | 1241 | 2.9% |
| Dodoma | 498 | 3.1% |
| Geita | 312 | 3.8% |
| Iringa | 289 | 4.2% |
| Kagera | 412 | 3.5% |
| Kaskazini Pemba | 145 | 6.8% |
| Kaskazini Unguja | 198 | 7.1% |
| Katavi | 198 | 3.9% |
| Kigoma | 341 | 3.2% |
| Kilimanjaro | 389 | 3.7% |
| Kusini Pemba | 145 | 6.4% |
| Kusini Unguja | 198 | 7.5% |
| Lindi | 241 | 2.4% |
| Manyara | 298 | 3.6% |
| Mara | 312 | 3.3% |
| Mbeya | 412 | 4.1% |
| Morogoro | 389 | 3.0% |
| Mtwara | 298 | 2.6% |
| Mwanza | 589 | 3.4% |
| Njombe | 198 | 4.4% |
| Pwani | 241 | 2.7% |
| Rukwa | 198 | 3.8% |
| Ruvuma | 241 | 0.9% |
| Shinyanga | 341 | 3.1% |
| Simiyu | 298 | 3.3% |
| Singida | 241 | 3.0% |
| Songwe | 198 | 3.7% |
| Tabora | 341 | 2.8% |
| Tanga | 389 | 3.2% |

| **Zone** | **Weighted n** | **CHW Contact %** |
| --- | --- | --- |
| Northern | 3421 | 22.4% |
| Central | 8912 | 22.8% |
| Southern | 8516 | 23.2% |

